# A pilot study to identify blood-based markers associated with response to treatment with Vedolizumab in patients with Inflammatory Bowel Disease

**DOI:** 10.1101/2024.09.19.24314034

**Authors:** John D. Rioux, Gabrielle Boucher, Anik Forest, Bertrand Bouchard, Lise Coderre, Caroline Daneault, Isabelle Robillard Frayne, Julie Thompson Legault, iGenoMed Consortium, Alain Bitton, Ashwin Ananthakrishnan, Sylvie Lesage, Ramnik J. Xavier, Christine Des Rosiers

## Abstract

The inflammatory bowel diseases (IBD) known as Crohn’s disease (CD) and ulcerative colitis (UC) are chronic inflammatory diseases of the gastrointestinal tract believed to arise because of an imbalance between the epithelial, immune and microbial systems. It has been shown that biological differences (genetic, epigenetic, microbial, environmental, etc.) exist between patients with IBD, with multiple risk factors been associated with disease susceptibility and IBD-related phenotypes (e.g. disease location). It is also known that there is heterogeneity in terms of response to therapy in patients with IBD, including to biological therapies that target very specific biological pathways (e.g. TNF-alpha signaling, IL-23R signaling, immune cell trafficking, etc.). It is hypothesized that the better the match between the biology targeted by these advanced therapies and the predominant disease-associated pathways at play in each patient will favor a beneficial response. The aim of this pilot study was to identify potential biological differences associated with differential treatment response to the anti α4β7 integrin therapy known as Vedolizumab. Our approach was to measure a broad range of analytes in the serum of patients prior to initiation of therapy and at the first clinical assessment visit, to identify potential markers of biological differences between patients at baseline and to see which biomarkers are most affected by treatment in responders. Our focus on early clinical response was to study the most proximal effects of therapy and to minimize confounders such as loss of response that occurs further distal to treatment initiation. Specifically, we performed targeted analyses of >150 proteins and metabolites, and untargeted analyses of >1100 lipid entities, in serum samples from 92 IBD patients (42 CD, 50 UC) immediately prior to initiation of therapy with vedolizumab (baseline samples) and at their first clinical assessment (14-week samples). We found lower levels of SDF-1a, but higher levels of PDGF-ββ, lactate, lysine, phenylalanine, branched chain amino acids, alanine, short/medium chain acylcarnitines, and triglycerides containing myristic acid in baseline serum samples of responders as compared to non-responders. We also observed an increase in serum levels of CXCL9 and citrate, as well as a decrease in IL-10, between baseline and week 14 samples. In addition, we observed that a group of metabolites and protein analytes was strongly associated with both treatment response and BMI status, although BMI status was not associated with treatment response.

## INTRODUCTION

The inflammatory bowel diseases (IBD) are chronic relapsing inflammatory diseases of the gastrointestinal tract that affect over one million Americans. The current “Therapeutic Pyramid” approach to IBD treatment aims to prescribe the mildest therapies first, and then for non-responders, progressively intensify to agents with greater therapeutic effect (e.g. biologics) but at significantly greater cost. Recently, there has been a shift in this treatment paradigm whereby biologics are being considered earlier (1, 2). Distinguishing in advance patients who will respond to biologics from non-responders would lead to improved therapeutic response rates, better outcomes for patients, decreased complications and adverse events, less tissue damage, and a decrease in societal costs. Currently, there are no reliable tools that enable us to match individual patients with the appropriate therapy. This will become an even greater problem with multiple new biologics and small molecules expected on the market in the coming years.

To meet this important challenge, numerous studies seeking biomarkers that predict response to advanced therapies in IBD have been performed using serum/plasma, fecal or biopsy samples. These studies support the need and feasibility of finding biomarkers associated with response to advanced therapies in IBD however, most restricted their analyses to a single biomarker type. For example, several SNPs are associated with non-response versus durable response to anti-TNF therapy (3, 4). Immune-assay-based stratification has also seen some success (5, 6). Functional assays using the agent or its target have also provided information regarding the likelihood of response (7). Biomarker discovery attempts have also been made with biopsies with good success for example high pre-treatment expression of oncostatin M was strongly associated with anti-TNF therapy failure (8, 9).

In the current study, we focus on the response to treatment with vedolizumab, as it has a mechanism of action quite distinct from other advanced therapies. Rather than block cytokine signaling, vedolizumab is believed to interfere with immune cell trafficking to the intestines. Specifically, vedolizumab is an antibody that is specific to α4β7 integrin and was designed to block its binding to MAdCAM-1, which is expressed on venular endothelium. While it was originally designed to block T-cell trafficking to the inflamed intestinal tissue, more recent studies suggest other potential mechanisms such as blocking the trafficking of pro-inflammatory macrophages and/or dendritic cells to the gut, or by skewing the macrophage population away from a pro-inflammatory phenotype to a wound healing phenotype (10, 11).

While beyond the scope of this pilot project, our ultimate objective is to develop a biomarker panel to facilitate therapeutic decisions in the clinic, we thus chose to prioritize blood-based biomarker discovery for the easiest integration within clinical practice. The cohort consisted of 42 CD and 50 UC patients where Vedolizumab had been administered intravenously at weeks 0, 2, 6 and 14, and serum samples collected at weeks 0 and 14 (12). Assessment of treatment response was performed using standardized clinical scores: Harvey Bradshaw Index (HBI) for patients with Crohn’s disease and the simple clinical colitis activity index (SCCAI) (13, 14). Serum samples were evaluated for the level of a targeted set of 49 cytokines, chemokines, and growth factors, many of which were previously shown to be elevated in the serum of patients with IBD and/or are genetically associated with disease risk (15). In addition, these serum samples were also evaluated for the levels of 10 organic acids (OA), 21 amino acids (AA) and 88 acylcarnitines (AC); all key metabolite classes implicated in immune cell activity and/or signaling. Moreover, we used an untargeted lipidomics approach that has previously been used to identify host and microbial metabolic functions associated with CD pathogenesis (16). These analyses enable two specific objectives: the first was to study the baseline samples to determine whether any of these biological markers were associated with response to therapy, and the second was to determine whether any changes in biomarker levels between weeks 0 and 14 were different between patients who experienced clinical response or remission compared to non-responders.

## MATERIALS AND METHODS

### Subjects and Sample Collection

Characteristics of patients included in the current study are described in **Table S1**. Patients were recruited at the Boston Massachusetts General Hospital (MGH) in 2014-2015 as part of a prospective cohort previously described (12). All patients consented to have their data and samples be part of the Prospective Registry in IBD Study at MGH (PRISM). CD and UC patients starting vedolizumab therapy were considered for inclusion. Vedolizumab was administered intravenously at weeks 0, 2, 6 and 14 at a dose of 300 mg. Patients who had completed the 3 infusion loading doses (weeks 0, 2 and 6) were included in the study. Disease was considered clinically active when HBI score > 4 for CD and SCCAI score > 2 for UC. Study outcome was achievement of either clinical response or remission at week 14. Response was defined as a decrease from baseline HBI ≥ 3 for CD and a decrease from baseline SCCAI ≥ 3 for UC. Remission was defined as HBI ≤ 4 and SCCAI ≤ 2. Blood was collected in non-fasting subjects at baseline and week 14 in SST tubes, ensuring rapid coagulation, which were processed within 24 hours of collection to collect the serum. Processed samples were stored at –80°C until the various analyses were performed in 2016. A review of electronic medical records was performed to obtain demographic and disease-related variables. Laboratory testing of serum samples, as described below, was performed in 2016. All experiments were performed in accordance with relevant guidelines and regulations. The Boston investigative team had access to information that could identify individual participants during and after data collection, whereas the Montreal team that performed the laboratory and statistical analyses only had access to deidentified data and samples. The study was approved by the Institutional Review Board of Partners Healthcare (protocol number 2004P001067/MGH) and the ethics committee of the Montreal Heart Institute, known as the *Comité d’éthique de la recherche et du développement des nouvelles technologies* (CERDNT; protocol number 2005-23; 05-813). Written informed consent was obtained for all subjects.

### Cytokine/chemokine measurements

The concentration of the 49 serum analytes were analyzed by quantitative cytokine assays using Bio-Plex Pro™ human cytokine standard 27-Plex, 21-Plex and sCD40L Singleplex assays (Bio-Rad Laboratories, California, USA) according to the pre-optimized protocol provided by the manufacturer. Briefly, for each assay, the standard curve samples were plated in duplicate and serum samples, some plated in duplicates, completed the 96-well plates. Samples were processed on different plates and dates, using a balanced design. The serum was diluted 1:4 in the buffer provided. Plates were labelled according to the manufacturer’s specifications and read by Bio-plex MAGPIX (Bio-Rad Laboratories, California, USA) at 525nm and 625nm and identified/quantified using the CCD imager. The complete list of analytes tested can be found in **Table S2.**

### QC Luminex data

Obvious outlier samples were removed before processing the data. Fluorescence Intensity (FI) distributions were normalized between plates using ComBat (library SVA, R 3.2.0) (17). Standard curves were fitted using 5 parameters logistic curves on FI pooled from all plates. The detection threshold was defined as the bottom value of the standard curve. Analytes with more than 50% samples having FI below the detection limit were removed from the analysis after verification that out-of-bound status was not associated with phenotype. Most remaining analytes had more than 75% of samples within the range of reliable quantification (recovery of standard concentration 70%-130%). One analyte, RANTES, had samples with FI above the upper limit of detection. Data quality was evaluated based on correlation of replicates and proportion of samples within range of quantification. To minimize impact of strong outliers, outliers were replaced by a value equivalent to 1.5 IQR from the lower and upper quartiles. The concentrations were then computed from the standard curve, with out-of-range values replaced by the minimum (or maximum) quantified concentration.

### Metabolic Profiling

Serum samples were obtained as frozen aliquots (-80°C) which had not been previously thawed (15). Following recommended guidelines (18), stratified randomization of samples was achieved according to potential confounding factors, namely age, sex, disease status and treatment, to minimize batch-dependent bias. Metabolites were identified according to m/z and retention time. For the targeted analyses, metabolites were quantified using internal standards (see **Table S3)**. MS quality controls (QCs) were performed by injecting “in-house” plasma pool at the beginning, the end and every 12 runs. Samples were processed on different batches, using a balanced design.

#### Targeted amino and organic acids analysis

Levels of 21 amino acids and 10 organic acids were measured by gas chromatography-mass spectrometry (GC-MS) using methods previously described (19) with some modifications. Serum (200 μl) was extracted with methanol and hydroxylamine and a mixture of internal standards was added. Derivatization was performed using N-methyl-N-tertbutyldimethylsilyltrifluoroacetamide. Samples were injected onto a GC-MS and operated in electronic ionization mode with helium as reagent gas. Data were expressed as concentrations.

#### Targeted acylcarnitine profiling

Extended profiling of 88 acylcarnitines (ACs) was performed by liquid chromatography coupled to tandem mass spectrometry (LC-MS) as described (20). Serum samples (100 μl) were spiked with methanol containing internal standards and were extracted consecutively with ethyl acetate, ether and methanol. The supernatants were combined, concentrated and injected onto an HPLC (20). Data were monitored in positive dynamic MRM mode (dMRM) for precursor ions of m/z 85 and processed by Mass Hunter QQQ quantitative software. Semi-quantitative analysis was performed for carnitine and each acylcarnitine by normalizing to the selected internal standard and data were expressed in signal intensity ratios. In the text, we used the LIPID MAPS nomenclature (ref: <u>LIPID MAPS</u>). Isomers of the same AC species, whose structure remains to be elucidated, are indicated using numbers (e.g., AC#1/#2).

#### QC of targeted metabolites

Obvious outlier samples were removed before processing the data. Missing values were presumed to be below detection range and thus, imputed to be 90% of the minimum detected value. To minimize the impact of strong outliers, outliers were investigated and replaced by a value equivalent to 2 IQR from the lower and upper quartiles.

#### Untargeted lipidomic profiling

Serum samples were analyzed using a previously validated semi-quantitative untargeted lipidomic workflow for plasma and serum (21) on a high-resolution LC-MS instruments (LC-quadrupole-time-of-flight (LC-QTOF 6530; Agilent Technologies Inc.). MS data were acquired and QC measures performed as recently described(16). Raw MS data were processed as previously described in detail using Mass Hunter Qualitative Analysis (version B.06 or B.07; Agilent, Santa Clara, USA) for peak picking and in-house bioinformatic scripts (available at MetaboICM/Metaborose (github.com) and MetaboICM/Data-processing: Script (github.com)) that for RT alignment, filtering, missing data imputation and batch correction(21). A maximal value of was set for the percentage of missing values or a given feature in any groups at 20%, and for coefficient of interindividual variation <80%. We had previously determined that median interindividual MS signal intensity variations in lipid features for the analysis of serum samples from four non-fasting subjects prepared using our standard optimal collection protocol was 33% and was manageable for successful application of our lipidomic workflow to serum samples collected in patient cohorts (16). The resulting final dataset, thereafter, referred to as features, defined by their m/z, RT and signal intensity. Lipid features of interest were then annotated to unique lipids with their respective acyl chains, by MS/MS analysis, as previously described (21).

### Statistical associations analyses

Statistical analyses were performed in R on log_2_ transformed data and thus represent fold changes (FC). Single marker analyses for association to phenotype were performed using linear regression including correction for gender, and with/without a correction for body mass index (BMI, log_2_ transformed) to assess robustness. The targeted metabolites analyses were also corrected for batch. The analyses were stratified for UC and CD. For lipidomics and proteomics, evidence of association for IBD was obtained by combining UC and CD results using inverse variance method. For targeted metabolites, the analysis was performed within all IBD. Disease activity was tested on all baseline samples. The response status was tested in active disease patients at baseline. From these patients with active disease at baseline, the subset with available data at week 14 was used to evaluate association between response status and markers ratio between week 14 and baseline. Multimarkers analyses were performed using logistic regression. Loess regression (R function loess, span=0.6) was used to represent association of BMI and specific markers.

For lipidomics, modules of correlated lipid features were created using the WGCNA (version, v1.64–1) package in R, on log_2_ transformed data. The type of network used was “signed”, with the “bicor” correlation function and softpower parameter set to 9. Minimal module size was set to 20 and hierarchical tree was cut at a height of 0.4 to determine clusters membership. We obtained 18 clusters, each represented by their respective first principal component.

Significant analytes were combined to evaluate their joint potential to predict response to therapy. All significant analytes from targeted analyses (14), together with the first principal component from lipidomics, were selected. Pairwise pearson correlation (r < 0.6) was used to reduce the number of parameters, after prioritization of the most associated analytes. A logistic regression analysis was performed, followed by stepwise bidirectional selection based on Aikake Information Criterion (AIC) was performed to reduce the model. The final model was evaluated for area under the roc curve (AUC). To be noted, no independent out of sample evaluation was performed, so it is expected that the performance might be overestimated.

## RESULTS

### Overall Study Design

Ninety-two patients were recruited at the Boston Massachusetts General Hospital (MGH) as part of a prospective cohort previously described (12). CD (n=42) and UC (n=50) patients starting vedolizumab therapy were recruited. The demographic and clinical phenotypes of these subjects are presented in **Table S1**. Serum samples were collected just prior to initiation of treatment with vedolizumab and at week 14. Statistical analyses were performed to examine two main objectives: (1) Identify which analytes had baseline levels that were different in responders as compared with “non responders”; (2) Identify analytes that changed significantly between baseline and after 14 weeks of treatment with vedolizumab in responders versus non-responders. In the former analysis, the intention is to identify biological pathways that favor treatment success in responders, and the latter analysis is aimed at identifying biological pathways that are modified by therapy in responders. In both analyses, “responder status” was defined as any patient having either clinical response or remission at week 14 (see Methods). These two analyses were performed only in patients with active disease at baseline. In addition, to place these analytes in context of disease activity, we also compared baseline levels in patients with active disease and those with quiescent disease. Finally, testing for association between each analyte and responder status was performed separately in CD and UC, as treatment patterns and outcomes are often different between these diseases. Nonetheless, given that multiple biological pathways are common to these two diseases we also tested for association in the combined set of CD and UC cases to search for potential commonalities. Although we highlight in the text below the phenotype (CD, UC, IBD) where the association was the most significant, the full set of analyses are presented in the accompanying tables.

### Targeted serum proteomics

As a first step to identify biomarkers that are associated with response to Vedolizumab treatment in IBD, we tested the serum samples from all 92 patients for 49 different analytes that represent products of genes within IBD loci, ligands for proteins encoded by genes within IBD loci, as well as a broad selection of cytokines, chemokines and growth factors known to play a role in a variety of immune functions (**Table S2**). Following quality control analyses, 142 samples from 90 individuals (52 at week 14) were included and 35 of the analytes were deemed detectable with reliable quantification in patient sera (**Table S2**). We first examined if baseline levels of these analytes were significantly associated (P<0.01) with response to treatment with vedolizumab. Indeed, we observed a nearly 2-fold greater baseline serum levels (P=7.0×10^-3^) of homodimeric platelet-derived growth factor subunit bb (PDGFßß) in UC patients responding to treatment as compared to non-responders (**Table 1**; **Fig. 1A**). Importantly, PDGF has been linked with angiogenesis in the context of chronic inflammatory processes as well as in gut wound healing (22). While less significant (P<0.05), there were four additional analytes (IL-1Ra, IL-4, IL-9, MIP-1b P<0.05) that were elevated at baseline in responders with all but IL-4 being associated with M1 macrophages, whereas CXCL12/SDF-1a, a chemokine associated with polarization of M2 macrophages (23), was 26% lower (FC=0.74, P=0.024) in responders (**Fig. 1B**). Interestingly, none of the analytes tested in baseline samples was significantly associated with disease activity (**Table 1, Table S2**). Steroid use at baseline was not associated with response to therapy and had negligeable impact on our results (**Table S2**). Amongst responders, five CD and nine UC were in steroid-free remission after 14 weeks of therapy. Interestingly, while lower baseline levels of CRP were found to be suggestively associated to response in IBD (P=0.077), the associations reported between proteomics and response are mostly independent from CRP (**Table S2**). In particular, in a logistic regression model for response including both log_2_(CRP) (OR=0.84, P= 0.31), and log_2_(PDFG-ßß) (OR=2.59, P= 0.049), only PDFG-ßß remained significant. Similarly, in a logistic regression model for response including both log_2_(CRP) (OR=0.87, P= 0.40), and log_2_(SDF1-a) (OR=0.19, P= 0.048), only SDF1-a remained significant.

**Figure 1.**
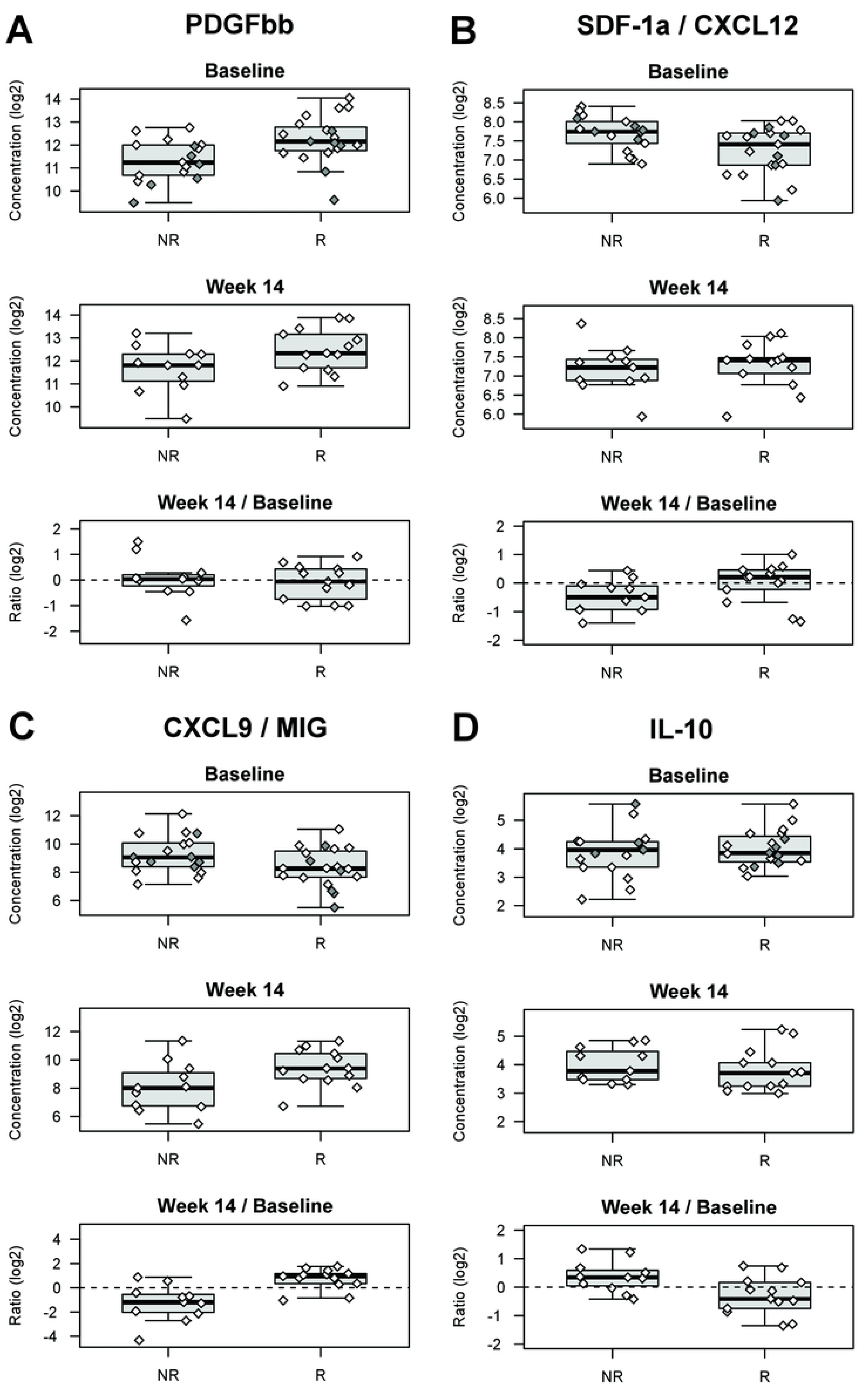
Targeted serum protein analyte levels associated with response to Vedolizumab treatment. Responders to vedolizumab treatment, had higher baseline serum levels of PDGFßß than non-responders **(A).** In contrast, responders had lower serum levels of SDF-1a at baseline as compared to non-responders **(B).** In responders, CXCL9/MIG levels increased 61% (FC=1.61) between baseline and week 14, whereas there was a 66% decrease (FC=0.44) in non-responders **(C).** In contrast, serum levels of IL-10 decreased 15% (FC=0.85) in responders, with a 27% increase (FC=1.27) in non-responders **(D).** Filled symbols are for individuals where samples were only available at baseline. All plots shown are for “UC”. See also **Table S2**.

**Table 1.**
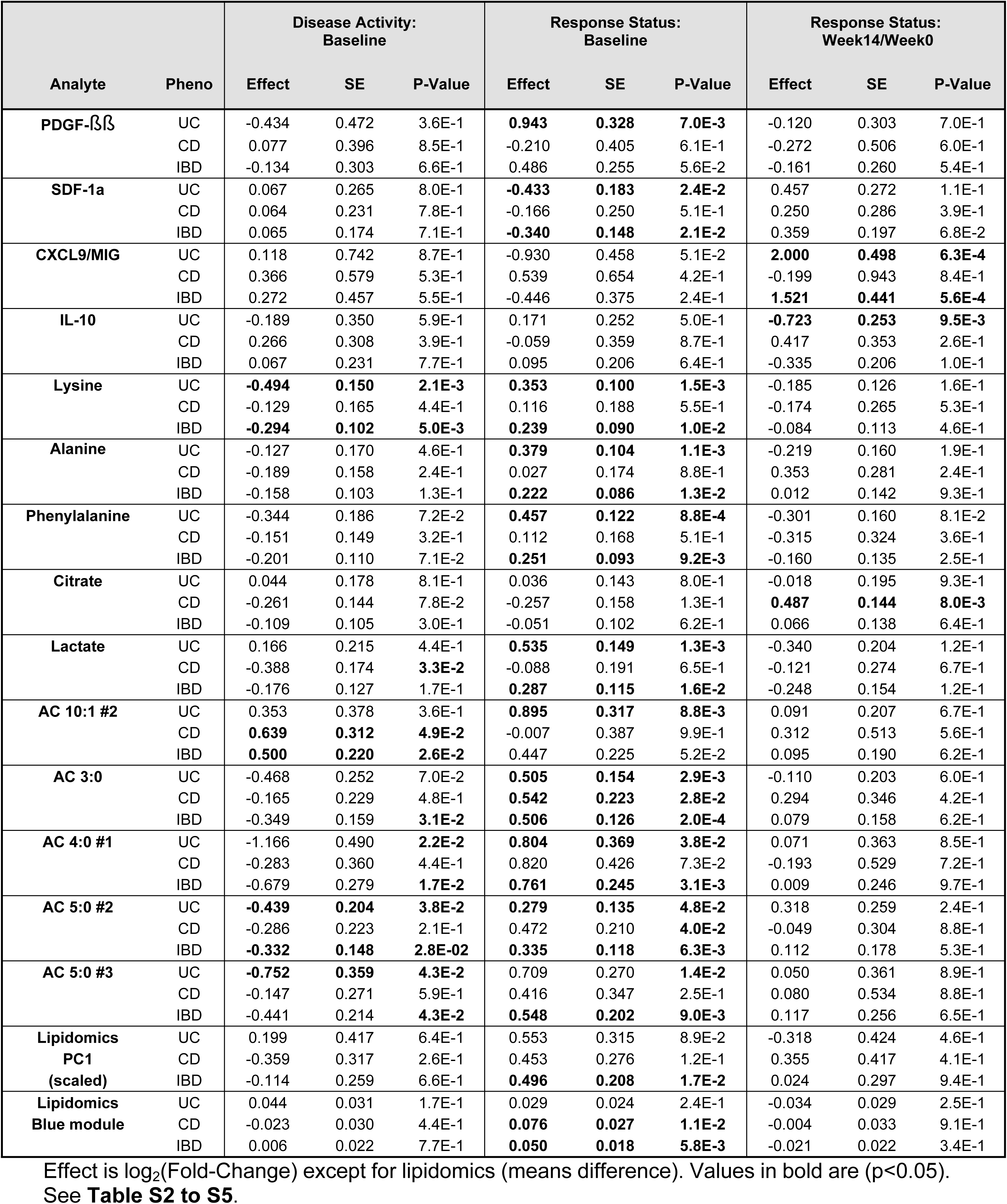
Top analytes associated with response to vedolizumab treatment.

Next, we assessed whether there were any changes in analyte levels between baseline and week 14 samples that were associated (P<0.01) with response to treatment with vedolizumab. The two strongest differences were for CXCL9/MIG (P=6.3×10^-4^), and for IL-10 (P=9.5×10^-3^) in patients with UC. Specifically, in responders CXCL9/MIG levels increased 61% (FC=1.61) between baseline and week 14, whereas there was a 66% decrease (FC=0.44) in non-responders (**Fig. 1C**). In contrast, serum levels of IL-10 decreased by 15% (FC=0.85) in responders, with a 27% increase (FC=1.27) in non-responders (**Fig 1D**). While not as statistically significant (P<0.05), we also observed an opposite effect of treatment on IL-1b, IL-12p70, FGF and G-CSF in responders (decrease) vs non-responders (increase). These results were found to be robust to steroid status changes between baseline and week 14. Changes in CRP levels were not associated with response (P=0.97).

### Targeted serum metabolomics – organic acids, amino acids, and acylcarnitines

Serum levels of 10 organic acids and 21 amino acids were measured by isotope dilution gas chromatography coupled with mass spectrometry (GC-MS). Regarding organic acids, we observed changes in common proxies of cell energy metabolism, namely in baseline samples, higher levels of lactate, reflecting cytosolic anaerobic glycolysis, (FC=1.45, P=1.3×10^-3^) that were associated with response to treatment in UC patients (**Fig. 2**) and between baseline and week 14 samples, a larger increase (FC=1.44, P=8.0×10^-3^) in levels of citrate, the first intermediate of mitochondrial Krebs cycle, in patients with CD responding to treatment, compared to non-responders. (**Fig. 2**).

**Figure 2.**
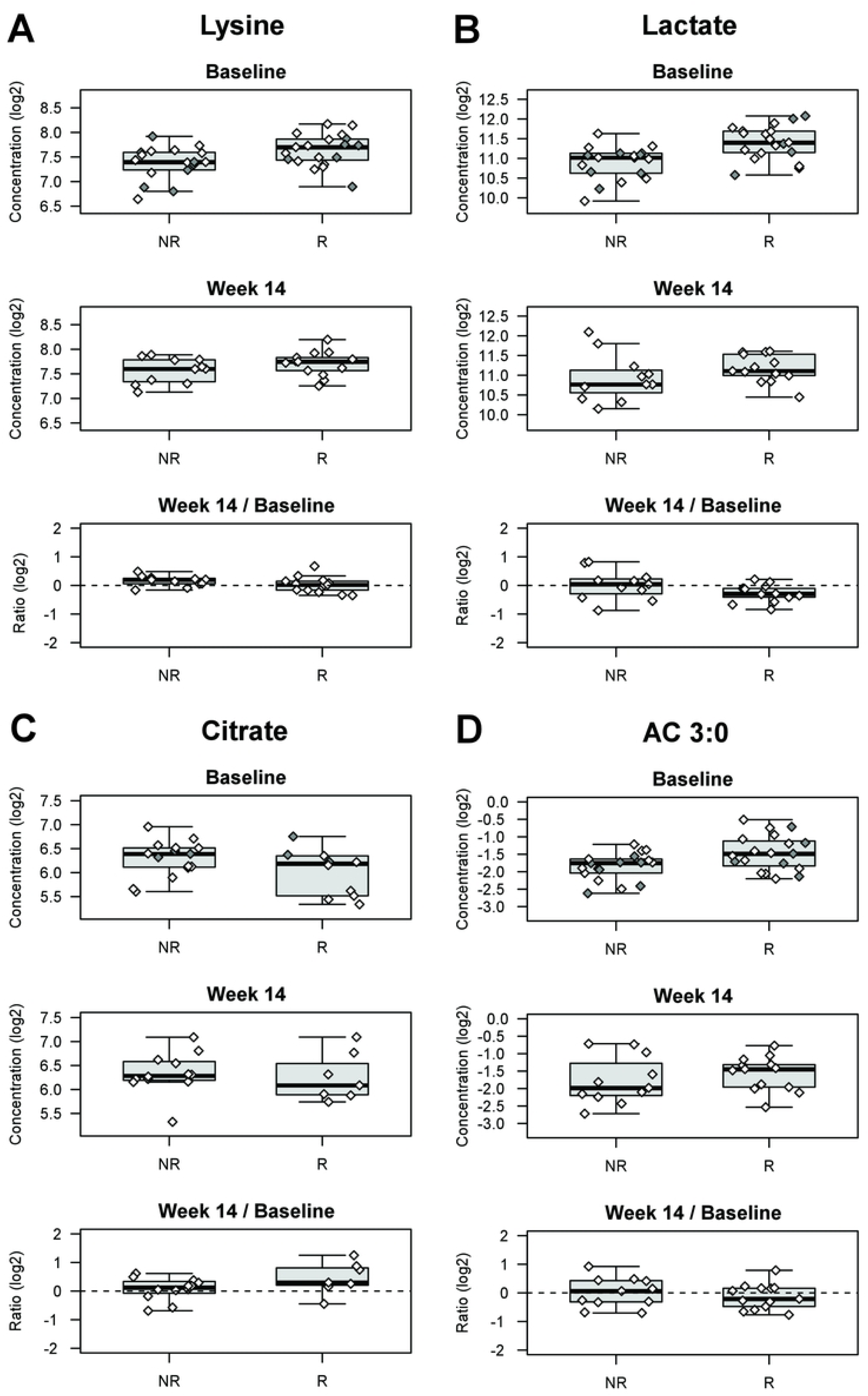
Targeted serum metabolite levels associated with response to Vedolizumab treatment. A selected set of analytes that were found to be associated with response at 14 weeks are presented. Elevated baseline levels of the amino acid lysine **(A)** and the organic acid lactate **(B)** were associated with response to treatment in patients with UC (P=2.1×10^-3^ and P=1.31×10^-3^, respectively). We also observed that ∼2-fold increase in serum levels of the organic acid citrate was associated with response in patients with CD **(C)**. Baseline serum levels of multiple acyl carnitines, including AC 3:0 were also associated (P=2.9×10^-3^) with response **(D)**. Filled symbols are for individuals where samples were only available at baseline. All plots shown are for “UC”, except for citrate (C), where the plot is shown for “CD”. See also **Table S3**.

As for amino acids, we observed that the baseline levels of multiple amino acids were significantly associated with therapeutic response (**Table 1; Table S3**). In particular, essential amino acids, namely the branched chain amino acid (BCAA) leucine (P=9.3×10^-3^), the sulfur-containing methionine (P=1.3×10^-3^), and phenylalanine (P=9.2×10^-3^), were significantly higher at baseline in IBD patients that responded to therapy as compared to non-responders. This was also observed in the UC subgroup, although many more amino acids were elevated in baseline samples of responders, such as lysine and alanine, the latter concurring with our findings of higher lactate levels (P<2×10^-3^) (**Fig. 2**; **Table S3**). Interestingly, when comparing baseline samples from patients with active disease to those with quiescent disease, we observed some overlap with the previous analysis and as well as a distinct set of metabolites associated with active disease. For example, lower serum levels of asparagine, glycine, histidine, and lysine were associated with active disease in UC patients (**Table S3**). We did not observe any significant changes in amino acid levels between baseline samples and those taken at 14 weeks post initiation of therapy.

Extended profiling of 88 ACs was performed by liquid chromatography coupled to tandem mass spectrometry. Acylcarnitines are recognized proxies of perturbations in fatty acid and BCAA oxidation, but can also exert biological activities, including to activate pro-inflammatory pathways in macrophages (24). In patients with IBD, both UC and CD, we found that elevated levels in baseline samples of four acylcarnitines containing short odd- and even chains (AC 3:0, AC 4:0 #1, AC 5:0 #2, AC 5:0 #3) were associated (P<0.01) with response to therapy (**Fig. 2**). Lower levels of these same acylcarnitines were modestly associated (P<0.05) with active disease suggesting that successful treatment normalized the levels of these metabolites. In patients with UC, response was also associated with elevated levels of AC 10:1 #2 in baseline samples. While there is a difference in specific identities between IBD and UC analyses, it should be noted that all are short-to-medium chain acylcarnitines. We did not detect any changes in acylcarnitine levels between baseline and week 14 samples that were associated with response in patients with CD, UC, or IBD.

In addition to analyzing individual metabolites, we also considered 11 ratios and groupings of metabolites (**Table S3**). For baseline values, grouping all odd short chains AC (AC 3:0, AC 5:1, AC 5:0 #1, AC 5:0 #2, AC 5:0 #3), associated with BCAA metabolism, was associated to response in IBD (P=8.4×10^-4^, FC=1.36), UC (P=4.6×10^-3^, FC=1.38) and suggestively in CD (P=0.06, FC=1.37). We also found evidence of association for BCAA (Leucine, Isoleucine, Valine) with response to therapy in UC (P=3.5×10^-3^, FC=1.33) and IBD (P=0.011, FC=1.21).

### Untargeted serum lipidomics

We conducted LC-MS-based lipidomic profiling on these same samples. Raw data consisting of 1364 MS signals were processed as previously described (16, 21) for RT alignment, filters of presence, normalization of signal intensities, and imputation of missing values. Following this data processing, the final dataset retained 1111 lipid features, defined by their m/z, RT and signal intensity. In IBD, we identified 63 lipid features that were significantly (P<0.05) elevated and 44 that were lower in baseline serum samples from responders as compared to non-responders (**Fig 3A**. Volcano plot). To account for multiple testing and because high dimensional lipidomic data is known to have a correlation structure, we used WGCNA and defined 18 clusters of correlated lipid features (illustrated as the colors in **Fig 3A**), each represented by their first principal component. We identified one cluster (denoted blue), associated with response to therapy in IBD (P=5.8×10^-3^) in baseline samples (**Table S4**). This module was also found to be associated with diagnosis (higher in CD vs UC, P=9.1×10^-3^). Next, we performed principal components analysis with all 1111 lipid features on baseline samples and observed that the first principal component (PC1) was also associated with response status in IBD (P=0.017) (**Fig. 3B**, **Table 1, Table S4**). Notably, the first principal component (PC1) was also significantly associated with body mass index (BMI) (P=5.1×10^-5^), while BMI was not found to be significantly associated to response status (P=0.68). Conditional analyses confirmed that the PC1 is associated to both BMI and response status, with a possible interaction between them. Overweight patients (BMI >25 kg/m2) who responded to therapy had higher values on PC1 at baseline than non-responders of similar BMI (**Fig. 3C**). When examining the loadings for the individual metabolites for PC1, we observed a predominant impact of lipid features from the same module (blue module) (**Fig 3D**), to which PC1 was also found to be correlated (r=0.77, P=8×10^-16^). Investigating on the nature of this signal, we identified at least six metabolites that were significantly correlated with PC1 (r>0.6, P<5×10^-8^), contributed a large amount of the component (loading > 7.5%) and that were significantly associated with response to therapy (P <0.01, q<0.20) (**Table S5** and **Fig. 3D**). These were all triglycerides (TG) and were elevated in baseline samples of responders in comparison with NR: TG 40:0 (P=2.9×10^-3^); TG 42:0 (P=3.2×10^-3^); TG 42:1 (P=7.5×10^-4^); TG 42:2 (P=3.1×10^-4^); TG 44:2 (P=1.0×10^-3^); TG 48:0 (P=2.7×10^-3^). Triglycerides are formed from glycerol and three fatty acids (FA). We thus performed MS/MS analyses to identify the specific fatty acids that constitute these TGs and all contained medium chain FA (MCFA), with 10 to 12 carbons in length, and/or long chain fatty acids (LCFA), with 14, 16, or 18 carbons in length; primarily saturated or monounsaturated (**Table 2, Table S5**). Interestingly, almost all contained myristic acid (C14). In terms of changes of individual lipid metabolites between baseline and week 14 samples in patients with CD, the PC1 and blue module described above were not found to be significantly impacted by therapy (P=0.89, P=0.18) and there was no significant difference between responders and non-responders. It is noteworthy that PC1 was not found to be associated with CRP levels (r=-0.02, P=0.87), suggesting that these are capturing different biological pathways.

**Figure 3.**
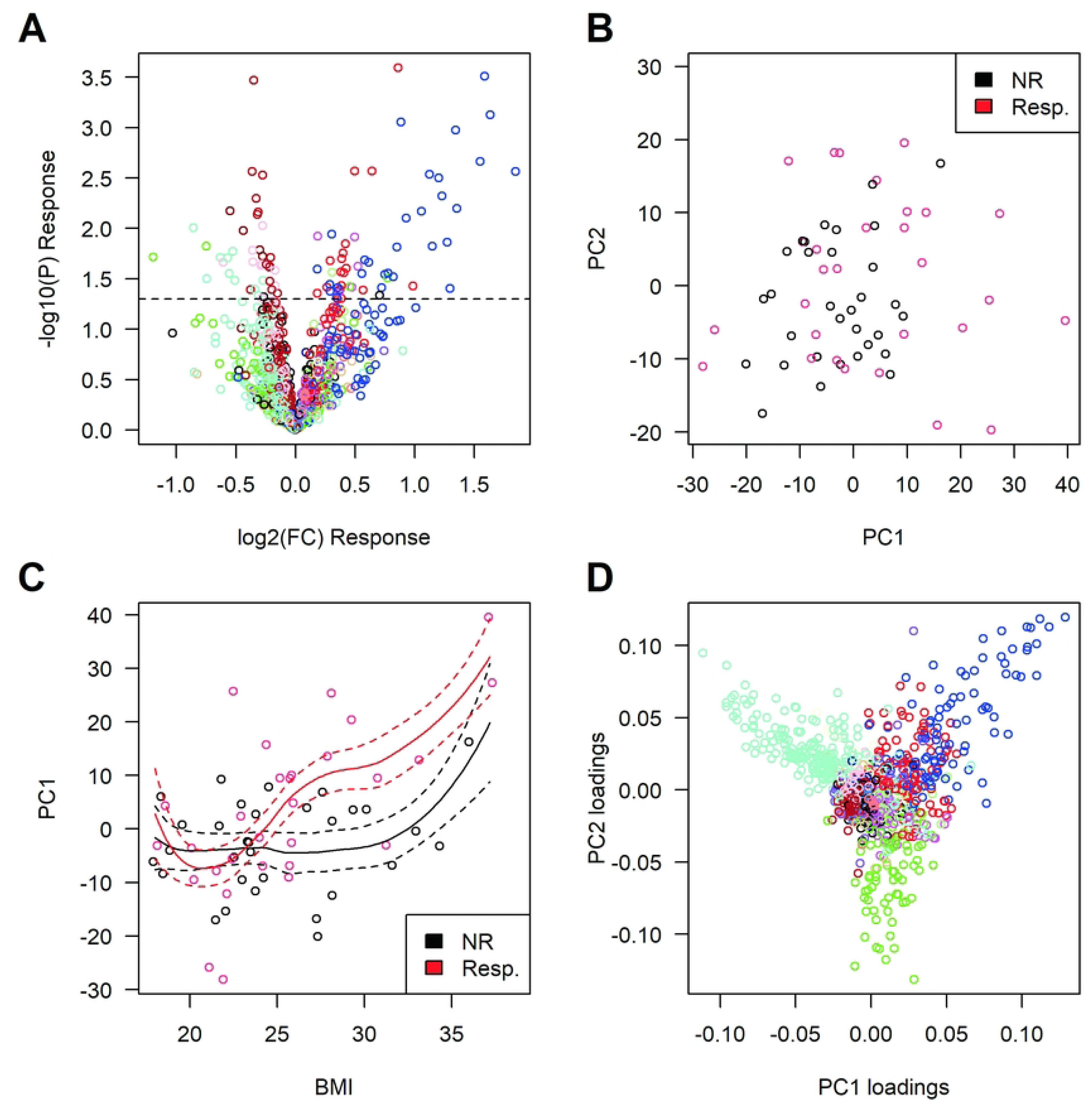
Untargeted serum LC-MS-based lipidomic analyses. We identified 107 lipid features that were significantly (P*<0.05*) associated to response (**A**). Colors in the volcano plot represent the 18 clusters of metabolites identified by WGCNA. The first component (PC1) from the PCA of the 1111 lipid features was significantly associated (P=0.017) with response (**B**). This PC1 was not only associated with response to therapy but was also associated with BMI (P=5.1×10^-5^) (**C**); dotted lines represent SE of loess fit. Importantly, we did not detect any significant association between BMI and response. The PCA loadings value for each lipid feature was plotted, illustrating that PC1 is primarily driven by the “blue” WGCNA cluster, which is also associated to response (P=5.8×10^-3^) (**D**). We also found an opposite impact for features from another module (named turquoise), which was not significantly associated to response to therapy (P=0.44). See also **Table S4** and **S5**.

**Table 2.** Top lipid features.

| Compound Name | Lipid abbreviation | Pheno | Response Status:<br>Baseline |  |  | BMI :<br>Baseline |  |  | Correlation with PC1 |  |  |
| --- | --- | --- | --- | --- | --- | --- | --- | --- | --- | --- | --- |
|  |  |  | log2(FC) | SE | P-Value | log2(FC) | SE | P-Value | Pearson<br>r | P-value | Loadings |
| POS:740.5911@41.37 | TG42:2 | UC | 1.41 | 0.56 | 1.6E-02 | 2.03 | 1.24 | 1.1E-01 | 0.64 | 6.9E-10 | 0.091 |
|  |  | CD | 1.87 | 0.72 | 1.7E-02 | 0.60 | 1.13 | 6.0E-01 |  |  |  |
|  |  | IBD | 1.59 | 0.44 | 3.1E-04 | 1.25 | 0.83 | 1.3E-01 |  |  |  |
| POS:711.6357@43.30 | TG40:0 | UC | 0.96 | 0.45 | 4.1E-02 | 2.85 | 1.00 | 7.8E-03 | 0.61 | 5.8E-09 | 0.076 |
|  |  | CD | 1.50 | 0.69 | 4.1E-02 | 1.73 | 1.08 | 1.2E-01 |  |  |  |
|  |  | IBD | 1.12 | 0.38 | 2.9E-03 | 2.34 | 0.74 | 1.5E-03 |  |  |  |
| POS:737.6516@43.56 | TG42:1 | UC | 1.27 | 0.57 | 3.4E-02 | 3.85 | 1.27 | 4.9E-03 | 0.65 | 5.1E-10 | 0.110 |
|  |  | CD | 2.57 | 0.91 | 1.1E-02 | 1.47 | 1.43 | 3.2E-01 |  |  |  |
|  |  | IBD | 1.64 | 0.49 | 7.5E-04 | 2.81 | 0.95 | 3.2E-03 |  |  |  |
| POS:763.6672@44.01 | TG44:2 | UC | 1.14 | 0.53 | 3.9E-02 | 3.60 | 1.17 | 4.4E-03 | 0.68 | 2.1E-11 | 0.099 |
|  |  | CD | 1.67 | 0.66 | 1.9E-02 | 1.42 | 1.03 | 1.8E-01 |  |  |  |
|  |  | IBD | 1.35 | 0.41 | 1.1E-03 | 2.37 | 0.77 | 2.2E-03 |  |  |  |
| POS:739.6673@46.76 | TG42:0 | UC | 0.97 | 0.49 | 5.5E-02 | 2.94 | 1.08 | 1.1E-02 | 0.64 | 8.9E-10 | 0.089 |
|  |  | CD | 1.75 | 0.75 | 2.9E-02 | 1.95 | 1.17 | 1.1E-01 |  |  |  |
|  |  | IBD | 1.20 | 0.41 | 3.2E-03 | 2.49 | 0.79 | 1.7E-03 |  |  |  |
| POS:823.7606@51.47 | TG44:0 | UC | 1.94 | 0.73 | 1.2E-02 | 4.66 | 1.61 | 7.0E-03 | 0.66 | 2.3E-10 | 0.129 |
|  |  | CD | 1.61 | 1.17 | 1.8E-01 | 1.40 | 1.83 | 4.5E-01 |  |  |  |
|  |  | IBD | 1.85 | 0.62 | 2.7E-03 | 3.23 | 1.21 | 7.5E-03 |  |  |  |

Loadings: loadings (rotation matrix) attributed to each lipid in the PCA for PC1.
See **Table S5** for further details.

In summary, the lipidomic PC1 and PC1-correlated analytes at baseline are associated to response to therapy and BMI, while BMI alone is not informative to predict response to therapy. There is also no evidence of an effect of therapy on these markers after 14 weeks.

### Correlation between biomarker types and logistic regression-based models

As it is likely that different biomarker types can reflect different aspects of the same pathway, we tested for correlation between the biomarkers reported above. Specifically, we looked for correlation between the lipidomic and proteomic data and found that the PC1 described above was found to be significantly correlated with CXCL1/GROa (r=-0.30, P=8.6×10^-3^), SDF-1a (r=-0.31, P=7.3×10^-3^) and IL-13 (r=0.32, P=4.7×10^-3^). As described above, lower baseline levels of SDF-1a were associated with response in IBD patients, while we found CXCL1/GROa to be suggestively associated (P=0.054). A logistic regression model including both lipidomic PC1 and SDF-1a explained more variation in response (P=8.8×10^-3^) than model including only the covariates (diagnosis and BMI). There is suggestive evidence that including both explains more variation than either one of them, pointing to a possible common signal that would be imperfectly captured by each parameter alone. In these models we obtained a similar, but slightly more significant association (P=3.3×10^-3^), when using the blue module instead of whole data PC1, suggesting that the observed association is primarily driven by lipids within this module.

The PC1 was also significantly correlated to many targeted metabolites, in particular to AC 3:0 (Pearson r=0.56, P=2.1×10^-7^), AC 4:0 (Pearson r=0.53, P=1.4×10^-6^) and the grouping of odd short chain AC (Pearson r=0.53, P=8.1×10^-7^) that we found to be associated to response to therapy in IBD (**Table S3**). Given their correlation to PC1, we then tested for association to BMI, which is what was observed (P<1E-3). Conditional regression identified that most of the signal is captured by AC 3:0, which remains associated with response to therapy when we condition on PC1, the other metabolites or SDF-1a. Investigating further on this, we identified for AC 3:0 a similar trend than PC1 for BMI and response to therapy, with overweight patients (BMI >25 kg/m2) who responded to therapy having higher values of AC 3:0 at baseline (**Fig S1**), suggesting that AC 3:0, PC1 and SDF-1a might be capturing a common biological state at baseline.

We then investigated the ability to predict response to therapy, using the most associated markers. Using AC 3:0 alone in the model for IBD gave an AUC=0.75. We then obtained a logistic regression model for predicting response based on PC1, AC 3:0, lactate, phenylalanine, AC 10:1 #2, PDGF-ßß, IL-1Ra, and SDF-1a. These were selected from all the associated analytes (p<0.05), after keeping only a subset with pairwise Pearson correlation < 0.6. The model was then reduced based on AIC (see Methods). The final model for IBD included AC 3:0, IL-1Ra and UC/CD phenotype. The AUC obtained was 0.78. The final model for UC included PDGF-ßß, lactate and AC 10:1 #2. The AUC obtained was 0.89. It is to be noted that these models are exploratory, as this process is subject to overestimation of performance

## DISCUSSION

The analyses of patient serum samples performed in this proof-of-concept study presented herein has identified candidate biomarkers associated with response to treatment with vedolizumab in patients with IBD. While these biomarkers must be validated in independent cohorts by other groups, these results support the notion that there are biological differences between responders and non-responders. While most of the strongest associations were observed in the UC subgroup, future studies will be required to determine whether this was simply due to sample size or biological differences. Given the known functions of the analytes and metabolites that were found associated with response to treatment in this cohort, we were interested in examining how these findings could be placed in the context of the known mechanism of action for vedolizumab. Specifically, it is believed that it interferes with leukocyte trafficking to the intestines (25).

The opposite changes in serum expression of CXCL9/MIG and IL-10 between baseline and week 14 samples is consistent with the former being pro-inflammatory chemokine, produced by macrophages, endothelial cells and fibroblasts (26), and the latter an anti-inflammatory cytokine, primarily produced by T helper cells, macrophages and dendritic cells (27, 28). One possible explanation is that successful treatment prevents CXCL9/MIG-producing pro-inflammatory cells from trafficking to the intestines, while not preventing the IL-10-producing cells from doing so. While the treatment with vedolizumab is likely to impact the trafficking of multiple different cell types expressing α4β7 integrin, including monocytes, macrophages, as well as B, T, and NK cells (29, 30), it is important to note that M1 macrophages are associated with expression of CXCL9/MIG and M2 macrophages with IL-10. Intriguingly, responders also had lower baseline levels of CXCL12/SDF-1a, a chemokine associated with M2 macrophages (23). In UC, we also found that elevated baseline serum levels of PDGFßß was associated with response to vedolizumab. It is believed that elevated levels of platelet derived growth factor (PDGF) may reflect active tissue remodeling given that it is a potent mitogen for cells of mesenchymal origin and a likely mediator of epithelial-mesenchyme transition (31).

Interestingly, the results from the targeted and untargeted metabolomics are also consistent with retention of M1 macrophages in the circulation, which contribute to type 1 immune responses, where IFN-y is the prototypical cytokine. Specifically, it is known that in pro-inflammatory M1 macrophages preferentially use the glycolytic pathway, whereas the anti-inflammatory M2 use fatty acid oxidation (FAO) and OXPHOS pathways for energy production (32, 33). Elevated levels of lactate, alanine and PC1 containing TG species in baseline samples are consistent with these metabolic differences (24, 34) . Furthermore, it has also been reported that macrophages activated with IFN-γ increase glucose uptake and lactate release, increases TG levels, and induces lipid droplet (LD) accumulation (35, 36). This LD accumulation is dependent on exogenous fatty acids and is characteristic of M1 macrophages (35, 36). Interestingly, the MS/MS analyses identified that most of these TGs had at least one myristic acid (C14) chain. Myristic acid is a side chain to Phorbol Myristate Acetate (PMA), which is a well know immune cell activator. Myristic acid alone can activate immune cells, and act as metabolic checkpoint for macrophage responses by promoting N-myristoylation (37–39).

While the hypothesis that we propose to explain our observations remains speculative, findings by Zeissig and colleagues, however, are consistent with our hypothesis as they observed that transcriptomic profiles that reflected lower M1 and higher M2 in the colonic tissue in response to Vedolizumab (11). This is also consistent with prior observations that an imbalance in M1/M2 promotes inflammation, with M1 macrophages dominating disease development, destructive aspects of inflammation, and disease severity in inflammatory diseases (40–42). Nonetheless, we can’t formally discount a metabolic shift from M2 to M1 macrophages within the circulation, or that the patterns of serum biomarkers observed are due to the retention of other cell types. In addition, this hypothesis does not exclude other mechanisms for differential response, for example, a recent study reported an attenuation of the gut associated lymphoid tissue in patients treated with Vedolizumab (43).

It should be acknowledged that these findings are based on a single cohort and thus need to be validated in independent cohorts, ideally within the context of prospective studies comparing multiple different advanced therapies to determine whether these observations are specific to vedolizumab treatment or are applicable to other molecularly targeted therapies. In addition, future studies would also include nutritional status of patients, measures of other clinical biomarkers, and potentially other measures of treatment response. In terms of CRP levels, we found some suggestive association between response and CRP at baseline, but none for CRP changes between baseline and week 14. To place these results in context, a review of 16 recent studies of response to Vedolizumab, association of CRP with response to vedolizumab was assessed in 7 of 16 studies; no association was detected in 4 studies, higher levels in responders in 2 studies, lower levels in responders in one study (11, 44–58). While the lack of endoscopy data in the current study can be viewed as a weakness of the current study, one could argue that clinical response/remission is more relevant outcome at this early timepoint of 14 weeks, and endoscopic changes more relevant to later time points, given a recent study that found that as ∼12% of patients achieve endoscopic remission at **week 26**, and ∼18% at **week 52** (59)). Finally, although these limitations highlight that the results of the current study will not immediately be translated into better treatment selection process, we believe that this study represents a proof-of-concept that a multi’omic approach can help to uncover the biological heterogeneity that is related to differential treatment response.

## Data Availability

Individual-level raw data will be made available via the NIDDK IBD Genetics Consortium (https://www.ibdgc.org/research/resources).

## ACKNOWLDGEMENTS

The members of the iGenoMed Consortium at the time of this study were (in alphabetical order): Alain Bitton^1*^, Gabrielle Boucher^2^, Guy Charron^2^, Christine Des Rosiers^2,3*^, Anik Forest^2^, Philippe Goyette^2^, Sabine Ivison^4^, Lawrence Joseph^5*^, Rita Kohen^1^, Jean Lachaine^6*^, Sylvie Lesage^3,7*^, Megan Levings^4*^, John D. Rioux^2,3*^, Julie Thompson Legault^2^, Luc Vachon^8^, Sophie Veilleux^9*^, Brian White-Guay^3*^. **Affiliations:** ^1^McGill University Health Centre, Montreal, Quebec; ^2^Montreal Heart Institute Research Center, Montreal, Quebec; ^3^Université de Montréal, Faculté de Médecine, Montreal; ^4^University of British Columbia, Vancouver; ^5^McGill University, Faculty of Medicine, Department of Epidemiology, Biostatistics and Occupational Health; ^6^Université de Montréal, Faculté de Pharmacie; ^7^Maisonneuve-Rosemont Hospital, Research Center, Montreal; ^8^LV Consulting, Montreal; ^9^Université de Laval, Québec. *Principal Investigators on grant # GPH-129341 with JDR as Leader and AB as co-Leader.

## AUTHOR CONTRIBUTIONS

John D. Rioux (Conceptualization: Lead; Validation: Lead; Resources: Equal; Writing – original draft: Lead; Supervision: Lead; Project administration: Lead; Funding acquisition: Lead; Writing – review & editing: Lead)

Gabrielle Boucher (Formal analysis: Equal; Data curation: Equal; Writing – original draft: Equal; Writing – review & editing Equal; Visualization: Equal)

Anik Forest (Formal analysis: Equal; Investigation: Equal; Data curation: Equal; Writing – review & editing: Equal; Visualization: Equal)

Bertrand Bouchard (Investigation: Equal)

Lise Coderre (Formal analysis: Supporting)

Caroline Daneault (Investigation: Equal)

Isabelle Robillard Frayne (Investigation: Equal)

Julie Thompson Legault (Writing – original draft: Equal; Writing – review & editing: Equal)

Alain Bitton (Funding acquisition: Lead)

Ashwin Ananthakrishnan (Resources: Equal)

Sylvie Lesage (Conceptualization: Supporting; Resources: Supporting)

Ramnik J. Xavier (Resources: Equal)

Christine Des Rosiers (Conceptualization: Lead; Validation: Lead; Resources: Equal,

Supervision: Lead, Project administration: Equal; Funding acquisition: Equal; Writing – review & editing: Equal)

## ABBREVIATIONS

AC: Acylcarnitines
AA: Amino acids
BMI: body mass index
BCAA: branched chain amino acid
FC: fold change
FI: fluorescence intensity
GC-MS: gas chromatography coupled with mass spectrometry
IQR: interquartile range
LC-MS: liquid chromatography coupled to tandem mass spectrometry
LCFA: long chain fatty acids
MCFA: medium chain FA
m/z: mass-to-charge ratios
OA: organic acids
PCA: principal components analysis
PC1: principal component 1 from lipidomics
QC: quality controls
RT: retention time
TG: triglycerides
WGCNA: Weighted Correlation Network Analysis

## REFERENCES

1. Lichtenstein GR, Loftus EV, Isaacs KL, Regueiro MD, Gerson LB, Sands BE. ACG Clinical Guideline: Management of Crohn’s Disease in Adults. Am J Gastroenterol. 2018;113(4):481–517.

2. Rubin DT, Ananthakrishnan AN, Siegel CA, Sauer BG, Long MD. ACG Clinical Guideline: Ulcerative Colitis in Adults. Am J Gastroenterol. 2019;114(3):384–413.

3. Barber GE, Yajnik V, Khalili H, Giallourakis C, Garber J, Xavier R, et al. Genetic Markers Predict Primary Non-Response and Durable Response To Anti-TNF Biologic Therapies in Crohn’s Disease. Am J Gastroenterol. 2016;111(12):1816–22.

4. Oliver J, Plant D, Webster AP, Barton A. Genetic and genomic markers of anti-TNF treatment response in rheumatoid arthritis. Biomark Med. 2015;9(6):499–512.

5. Baird AC, Mallon D, Radford-Smith G, Boyer J, Piche T, Prescott SL, et al. Dysregulation of innate immunity in ulcerative colitis patients who fail anti-tumor necrosis factor therapy. World J Gastroenterol. 2016;22(41):9104–16.

6. Fuchs F, Schillinger D, Atreya R, Hirschmann S, Fischer S, Neufert C, et al. Clinical Response to Vedolizumab in Ulcerative Colitis Patients Is Associated with Changes in Integrin Expression Profiles. Front Immunol. 2017;8:764.

7. Van den Brande JM, Koehler TC, Zelinkova Z, Bennink RJ, te Velde AA, ten Cate FJ, et al. Prediction of antitumour necrosis factor clinical efficacy by real-time visualisation of apoptosis in patients with Crohn’s disease. Gut. 2007;56(4):509–17.

8. Arijs I, Li K, Toedter G, Quintens R, Van Lommel L, Van Steen K, et al. Mucosal gene signatures to predict response to infliximab in patients with ulcerative colitis. Gut. 2009;58(12):1612–9.

9. Tew GW, Hackney JA, Gibbons D, Lamb CA, Luca D, Egen JG, et al. Association Between Response to Etrolizumab and Expression of Integrin alphaE and Granzyme A in Colon Biopsies of Patients With Ulcerative Colitis. Gastroenterology. 2016;150(2):477–87 e9.

10. Sly LM, McKay DM. Macrophage immunotherapy: overcoming impediments to realize promise. Trends Immunol. 2022;43(12):959–68.

11. Zeissig S, Rosati E, Dowds CM, Aden K, Bethge J, Schulte B, et al. Vedolizumab is associated with changes in innate rather than adaptive immunity in patients with inflammatory bowel disease. Gut. 2019;68(1):25–39.

12. Shelton E, Allegretti JR, Stevens B, Lucci M, Khalili H, Nguyen DD, et al. Efficacy of Vedolizumab as Induction Therapy in Refractory IBD Patients: A Multicenter Cohort. Inflamm Bowel Dis. 2015;21(12):2879–85.

13. Harvey RF, Bradshaw JM. A simple index of Crohn’s-disease activity. Lancet. 1980;1(8167):514.

14. Walmsley RS, Ayres RC, Pounder RE, Allan RN. A simple clinical colitis activity index. Gut. 1998;43(1):29–32.

15. Boucher G, Paradis A, Chabot-Roy G, Coderre L, Hillhouse EE, Bitton A, et al. Serum Analyte Profiles Associated With Crohn’s Disease and Disease Location. Inflamm Bowel Dis. 2021.

16. Ferru-Clement R, Boucher G, Forest A, Bouchard B, Bitton A, Lesage S, et al. Serum Lipidomic Screen Identifies Key Metabolites, Pathways, and Disease Classifiers in Crohn’s Disease. Inflamm Bowel Dis. 2023;29(7):1024–37.

17. Johnson WE, Li C, Rabinovic A. Adjusting batch effects in microarray expression data using empirical Bayes methods. Biostatistics. 2007;8(1):118–27.

18. Burla B, Arita M, Arita M, Bendt AK, Cazenave-Gassiot A, Dennis EA, et al. MS-based lipidomics of human blood plasma: a community-initiated position paper to develop accepted guidelines. J Lipid Res. 2018;59(10):2001–17.

19. Thompson Legault J, Strittmatter L, Tardif J, Sharma R, Tremblay-Vaillancourt V, Aubut C, et al. A Metabolic Signature of Mitochondrial Dysfunction Revealed through a Monogenic Form of Leigh Syndrome. Cell Rep. 2015;13(5):981–9.

20. Ruiz M, Labarthe F, Fortier A, Bouchard B, Thompson Legault J, Bolduc V, et al. Circulating Acylcarnitine Profile in Human Heart Failure: A Surrogate of Fatty Acid Metabolic Dysregulation in Mitochondria and Beyond. Am J Physiol Heart Circ Physiol. 2017:ajpheart 00820 2016.

21. Forest A, Ruiz M, Bouchard B, Boucher G, Gingras O, Daneault C, et al. Comprehensive and Reproducible Untargeted Lipidomic Workflow Using LC-QTOF Validated for Human Plasma Analysis. J Proteome Res. 2018;17(11):3657–70.

22. Alkim C, Alkim H, Koksal AR, Boga S, Sen I. Angiogenesis in Inflammatory Bowel Disease. Int J Inflam. 2015;2015:970890.

23. Mantovani A, Biswas SK, Galdiero MR, Sica A, Locati M. Macrophage plasticity and polarization in tissue repair and remodelling. J Pathol. 2013;229(2):176–85.

24. Rutkowsky JM, Knotts TA, Ono-Moore KD, McCoin CS, Huang S, Schneider D, et al. Acylcarnitines activate proinflammatory signaling pathways. Am J Physiol Endocrinol Metab. 2014;306(12):E1378–87.

25. Feagan BG, Rutgeerts P, Sands BE, Hanauer S, Colombel JF, Sandborn WJ, et al. Vedolizumab as induction and maintenance therapy for ulcerative colitis. N Engl J Med. 2013;369(8):699–710.

26. Ohmori Y, Schreiber RD, Hamilton TA. Synergy between interferon-gamma and tumor necrosis factor-alpha in transcriptional activation is mediated by cooperation between signal transducer and activator of transcription 1 and nuclear factor kappaB. J Biol Chem. 1997;272(23):14899–907.

27. Iyer SS, Cheng G. Role of interleukin 10 transcriptional regulation in inflammation and autoimmune disease. Crit Rev Immunol. 2012;32(1):23–63.

28. Italiani P, Boraschi D. From Monocytes to M1/M2 Macrophages: Phenotypical vs. Functional Differentiation. Front Immunol. 2014;5:514.

29. Tyler CJ, Guzman M, Lundborg LR, Yeasmin S, Zgajnar N, Jedlicka P, et al. Antibody secreting cells are critically dependent on integrin alpha4beta7/MAdCAM-1 for intestinal recruitment and control of the microbiota during chronic colitis. Mucosal Immunol. 2022;15(1):109–19.

30. Schleier L, Wiendl M, Heidbreder K, Binder MT, Atreya R, Rath T, et al. Non-classical monocyte homing to the gut via alpha4beta7 integrin mediates macrophage-dependent intestinal wound healing. Gut. 2020;69(2):252–63.

31. Wu Q, Hou X, Xia J, Qian X, Miele L, Sarkar FH, et al. Emerging roles of PDGF-D in EMT progression during tumorigenesis. Cancer Treat Rev. 2013;39(6):640–6.

32. Huang SC, Everts B, Ivanova Y, O’Sullivan D, Nascimento M, Smith AM, et al. Cell-intrinsic lysosomal lipolysis is essential for alternative activation of macrophages. Nat Immunol. 2014;15(9):846–55.

33. Kelly B, Pearce EL. Amino Assets: How Amino Acids Support Immunity. Cell Metab. 2020;32(2):154–75.

34. Li S, Gao D, Jiang Y. Function, Detection and Alteration of Acylcarnitine Metabolism in Hepatocellular Carcinoma. Metabolites. 2019;9(2).

35. Rosas-Ballina M, Guan XL, Schmidt A, Bumann D. Classical Activation of Macrophages Leads to Lipid Droplet Formation Without de novo Fatty Acid Synthesis. Front Immunol. 2020;11:131.

36. Singh A, Sen P. Lipid droplet: A functionally active organelle in monocyte to macrophage differentiation and its inflammatory properties. Biochim Biophys Acta Mol Cell Biol Lipids. 2021;1866(10):158981.

37. Aderem AA, Keum MM, Pure E, Cohn ZA. Bacterial lipopolysaccharides, phorbol myristate acetate, and zymosan induce the myristoylation of specific macrophage proteins. Proceedings of the National Academy of Sciences of the United States of America. 1986;83(16):5817–21.

38. Jia M, Wang Y, Wang J, Qin D, Wang M, Chai L, et al. Myristic acid as a checkpoint to regulate STING-dependent autophagy and interferon responses by promoting N-myristoylation. Nat Commun. 2023;14(1):660.

39. Tada M, Ichiishi E, Saito R, Emoto N, Niwano Y, Kohno M. Myristic Acid, A Side Chain of Phorbol Myristate Acetate (PMA), Can Activate Human Polymorphonuclear Leukocytes to Produce Oxygen Radicals More Potently than PMA. J Clin Biochem Nutr. 2009;45(3):309–14.

40. Liu B, Zhang M, Zhao J, Zheng M, Yang H. Imbalance of M1/M2 macrophages is linked to severity level of knee osteoarthritis. Exp Ther Med. 2018;16(6):5009–14.

41. Xia T, Fu S, Yang R, Yang K, Lei W, Yang Y, et al. Advances in the study of macrophage polarization in inflammatory immune skin diseases. J Inflamm (Lond). 2023;20(1):33.

42. Zhu W, Yu J, Nie Y, Shi X, Liu Y, Li F, et al. Disequilibrium of M1 and M2 macrophages correlates with the development of experimental inflammatory bowel diseases. Immunol Invest. 2014;43(7):638–52.

43. Canales-Herrerias P, Uzzan M, Seki A, Czepielewski RS, Verstockt B, Livanos AE, et al. Gut-associated lymphoid tissue attrition associates with response to anti-alpha4beta7 therapy in ulcerative colitis. Sci Immunol. 2024;9(94):eadg7549.

44. Boden EK, Shows DM, Chiorean MV, Lord JD. Identification of Candidate Biomarkers Associated with Response to Vedolizumab in Inflammatory Bowel Disease. Dig Dis Sci. 2018;63(9):2419–29.

45. Soendergaard C, Seidelin JB, Steenholdt C, Nielsen OH. Putative biomarkers of vedolizumab resistance and underlying inflammatory pathways involved in IBD. BMJ Open Gastroenterol. 2018;5(1):e000208.

46. Verstockt B, Verstockt S, Veny M, Dehairs J, Arnauts K, Van Assche G, et al. Expression Levels of 4 Genes in Colon Tissue Might Be Used to Predict Which Patients Will Enter Endoscopic Remission After Vedolizumab Therapy for Inflammatory Bowel Diseases. Clin Gastroenterol Hepatol. 2020;18(5):1142–51 e10.

47. Pauwels RWM, de Vries AC, van der Woude CJ. Fecal calprotectin is a reliable marker of endoscopic response to vedolizumab therapy: A simple algorithm for clinical practice. J Gastroenterol Hepatol. 2020;35(11):1893–901.

48. Allner C, Melde M, Becker E, Fuchs F, Muhl L, Klenske E, et al. Baseline levels of dynamic CD4(+) T cell adhesion to MAdCAM-1 correlate with clinical response to vedolizumab treatment in ulcerative colitis: a cohort study. BMC Gastroenterol. 2020;20(1):103.

49. Osterman MT, VanDussen KL, Gordon IO, Davis EM, Li K, Simpson K, et al. Epithelial Cell Biomarkers Are Predictive of Response to Biologic Agents in Crohn’s Disease. Inflamm Bowel Dis. 2021;27(5):677–85.

50. Holmer AK, Battat R, Dulai PS, Vande Casteele N, Nguyen N, Jain A, et al. Biomarkers are associated with clinical and endoscopic outcomes with vedolizumab treatment in Crohn’s disease. Therap Adv Gastroenterol. 2020;13:1756284820971214.

51. Bertani L, Barberio B, Fornili M, Antonioli L, Zanzi F, Casadei C, et al. Serum oncostatin M predicts mucosal healing in patients with inflammatory bowel diseases treated with anti-TNF, but not vedolizumab. Dig Liver Dis. 2022;54(10):1367–73.

52. Gonzalez-Vivo M, Lund Tiirikainen MK, Andreu M, Fernandez-Clotet A, Lopez-Garcia A, Murciano Gonzalo F, et al. Memory T Cell Subpopulations as Early Predictors of Remission to Vedolizumab in Ulcerative Colitis. Front Med (Lausanne). 2022;9:837294.

53. Gabriels RY, Bourgonje AR, von Martels JZH, Blokzijl T, Weersma RK, Galinsky K, et al. Mucosal Eosinophil Abundance in Non-Inflamed Colonic Tissue Is Associated with Response to Vedolizumab Induction Therapy in Inflammatory Bowel Disease. J Clin Med. 2022;11(14).

54. Abreu MT, Davies JM, Quintero MA, Delmas A, Diaz S, Martinez CD, et al. Transcriptional Behavior of Regulatory T Cells Predicts IBD Patient Responses to Vedolizumab Therapy. Inflamm Bowel Dis. 2022;28(12):1800–12.

55. Haglund S, Soderman J, Almer S. Differences in Whole-Blood Transcriptional Profiles in Inflammatory Bowel Disease Patients Responding to Vedolizumab Compared with Non-Responders. Int J Mol Sci. 2023;24(6).

56. Liu J, Fang H, Hong N, Lv C, Zhu Q, Feng Y, et al. Gut Microbiome and Metabonomic Profile Predict Early Remission to Anti-Integrin Therapy in Patients with Moderate to Severe Ulcerative Colitis. Microbiol Spectr. 2023;11(3):e0145723.

57. Roosenboom B, Wahab PJ, Smids C, Meijer J, Kemperman L, Groenen MJM, et al. Mucosal alpha4beta7+ Lymphocytes and MAdCAM+ Venules Predict Response to Vedolizumab in Ulcerative Colitis. Inflamm Bowel Dis. 2024;30(6):930–8.

58. Kim MK, Jo SI, Kim SY, Lim H, Kang HS, Moon SH, et al. PD-1-positive cells contribute to the diagnosis of inflammatory bowel disease and can aid in predicting response to vedolizumab. Sci Rep. 2023;13(1):21329.

59. Danese S, Sandborn WJ, Colombel JF, Vermeire S, Glover SC, Rimola J, et al. Endoscopic, Radiologic, and Histologic Healing With Vedolizumab in Patients With Active Crohn’s Disease. Gastroenterology. 2019;157(4):1007–18 e7.

